## Supplementary File for "Traces of pubertal brain development and health revealed through domain adapted brain network fusion"

Address for correspondence:

### Supporting Information

#### Supplementary Figures:

Supplementary Fig. 1: Correlation between raw features and first brain embedding for HBN

Supplementary Fig. 2: Associations between brain embeddings and puberty, both in cross-sectional and longitudinal data of the ABCD cohort

Supplementary Fig. 3: Distribution of Δ brain embedding in the ABCD sample stratified for sex and pubertal categories derived from caregivers` reports at 1 year follow-up.

Supplementary Fig. 4: Associations between brain embeddings and dimensional psychopathology, both in cross-sectional and longitudinal data of the ABCD cohort

Supplementary Fig. 5: Distribution of puberty ratings in the ABCD sample stratified for sex, timepoints, and respondent

Supplementary Fig. 6: Distribution of psychopathology measures in the HBN sample as total counts and percentages stratified for sex

#### Supplementary Tables:

Supplementary Table 1:  Association results for ABCD models

Supplementary Table 2: Association results for HBN models

Supplementary Table 3: Pubertal category conversion scheme

Supplementary Table 4: Frequencies of pubertal categories in the ABCD sample.

Supplementary Table 5: Frequencies of psychopathology measures in the HBN sample.

### Supplementary Figures

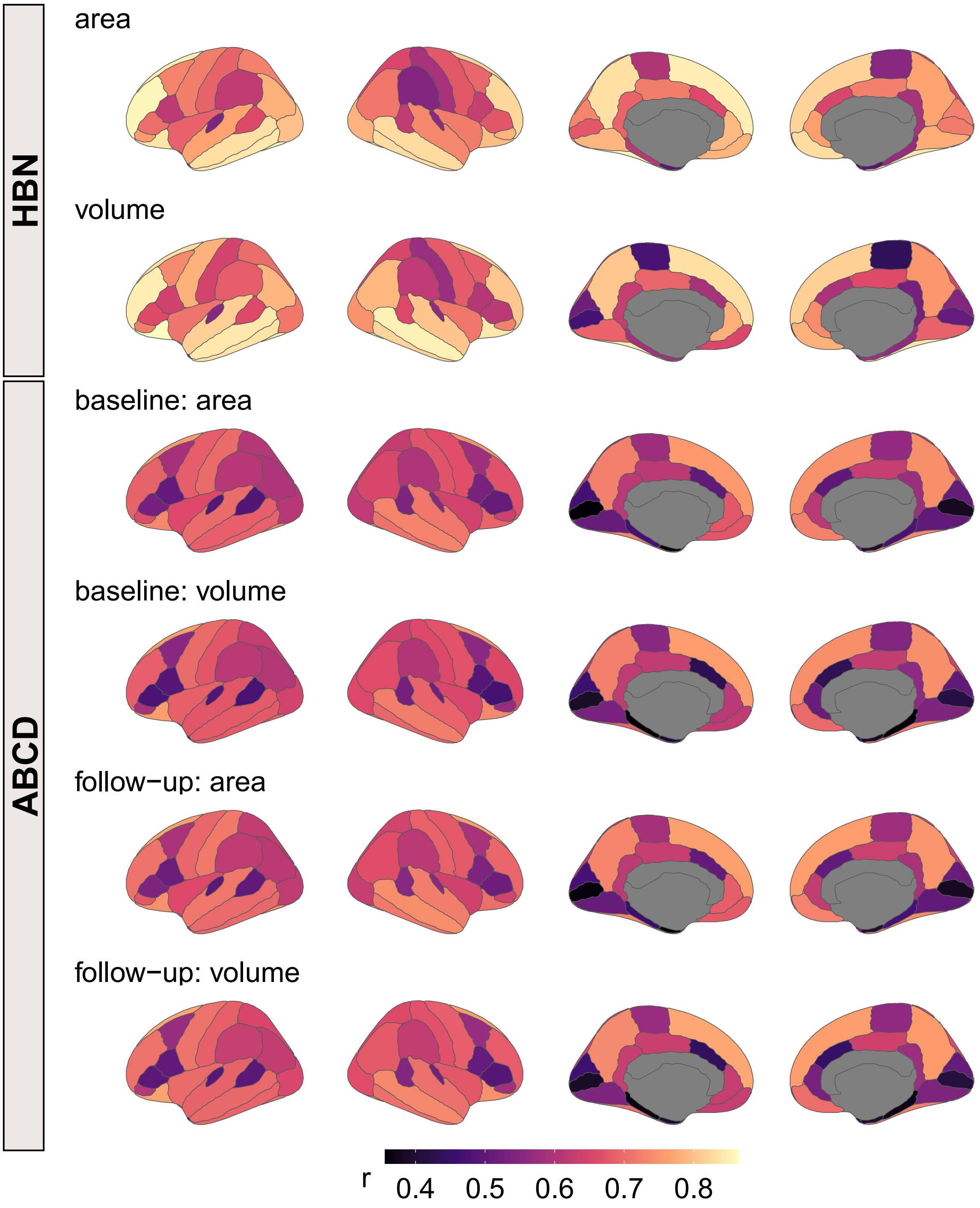

Supplementary Fig. 1: Correlation between raw brain features and first brain embedding. Correlation values indicate Pearson correlation coefficients. Visualization was performed with ggseg (version 1.6.5 [1]) in R (version 4.2.3). Since algebraic signs of the embeddings might be positive or negative (i.e., akin to an eigenvector), we multiplied negative correlations by (-1) to derive at comparable patterns across cohorts. Spin permutation testing with 5000 iterations indicated significant correlations between brain maps of the ABCD and HBN dataset (all $r > 0.64$, all $p < 0.0004$). Permutations were performed using the ENIGMA Toolbox (version 2.0.3, [2]).

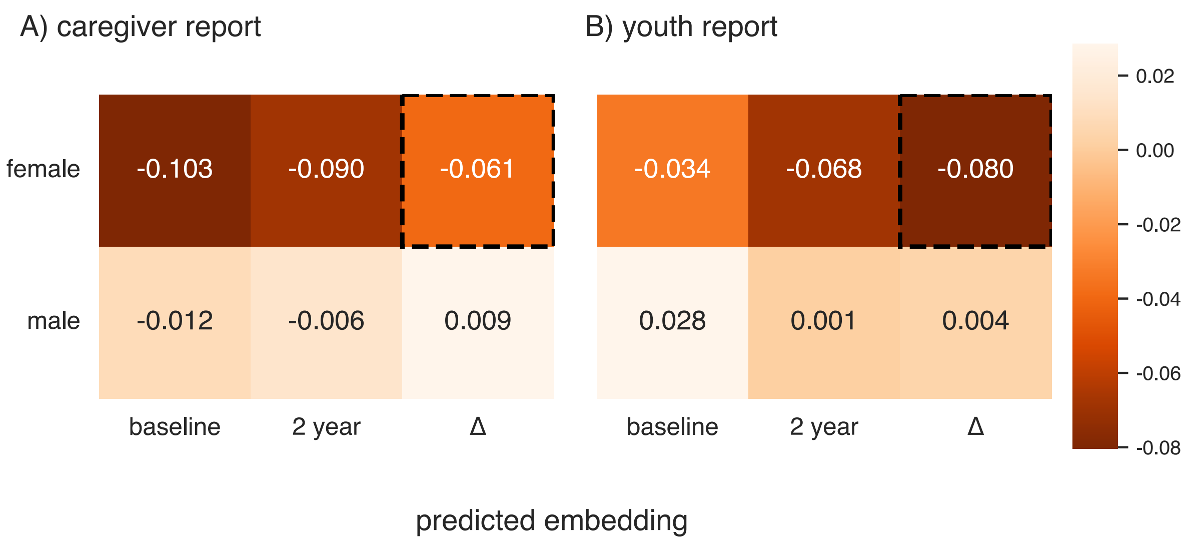

Supplementary Fig. 2: Associations between brain embeddings and puberty, both in cross-sectional and longitudinal data of the ABCD cohort. First two columns in A) and B) refer to associations between predicted brain embeddings and the respective pubertal score (PDS mean) per timepoint. Δ refers to the association between the Δ brain embedding and the ΔPDS mean score. Annotations refer to effect sizes from the linear models and highlighted cells with dashed lines indicate significant results. Linear models were performed in similar vein to the puberty models in the main manuscript but were expanded with Body Mass Index (BMI), Socioeconomic Status (SES), and race/ethnicity as covariates: we ran two cross-sectional models with age, SES, BMI, and race/ethnicity as covariates. Of note for SES there was only baseline data available. Additionally, we ran a longitudinal model in which the delta measures of the brain embedding, the change PDS score and the age and BMI difference between both timepoints were integrated. Moreover, baseline SES and race/ethnicity was added as covariate without a change score. BMI= (height (lb) / height (in)) x 703. Height and weight measures were averaged across two measurements. BMI smaller than 10 or higher than 50 were excluded after manual inspection. Race/ethnicity (ABCD variable acspsw03) refers to 5 different levels: White, Black, Hispanic, Asian, other. Other includes individuals identifying as multiracial and/or belonging to a race/ethnicity with too few members in the sample. For SES variables (i.e., parental education and total family income), “refuse to answer” and “don't know” where encoded as “NaN”. SES variables were transformed via rank-based inverse normal transformation and averaged resulting in a single SES status variable. Exact p-values and effect sizes can be found in Supplementary Table 1.

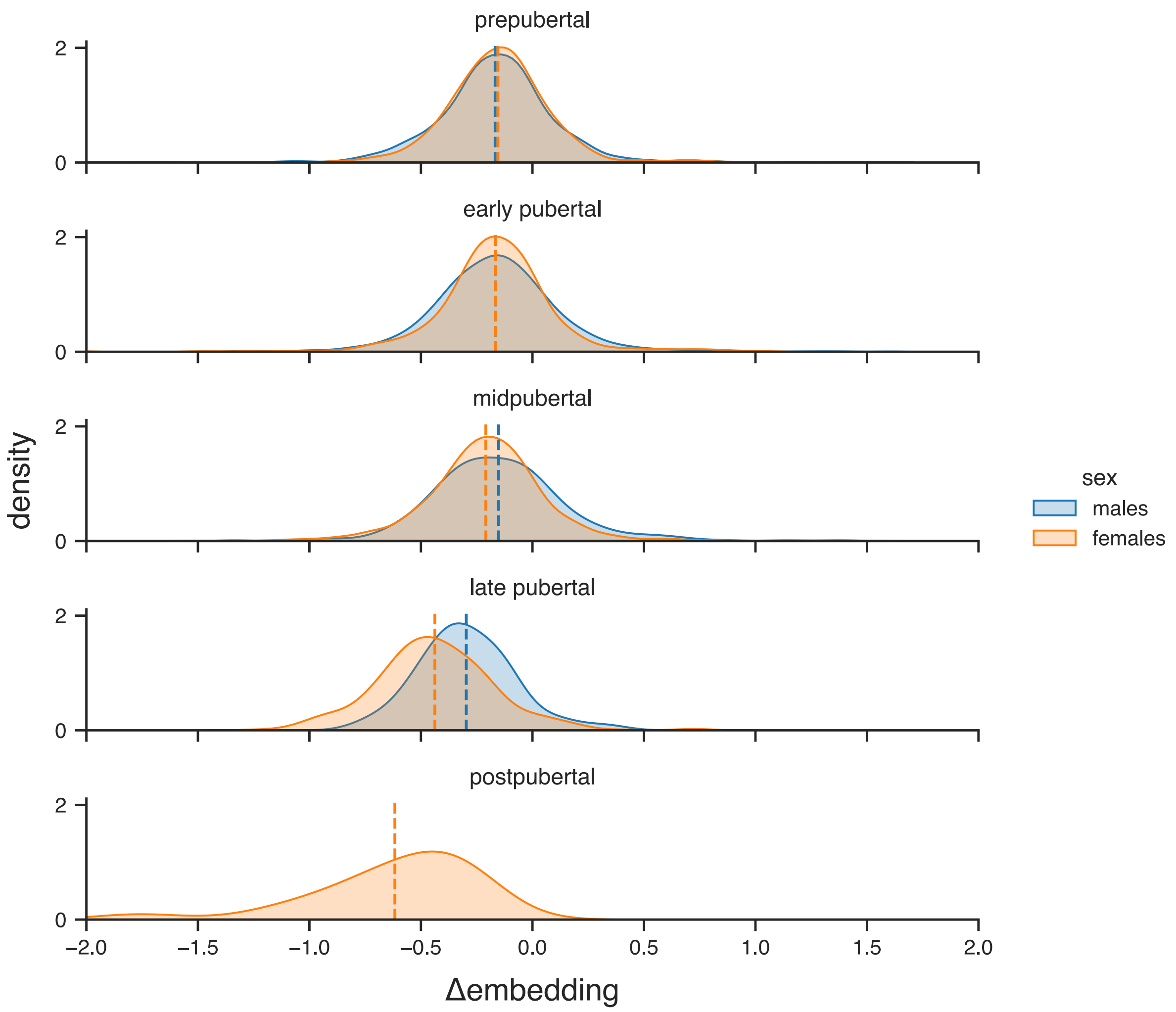

Supplementary Fig. 3: Distribution of Δ brain embedding in the ABCD sample stratified for sex and pubertal categories derived from caregivers` reports at 1 year follow-up. The figure shows the same pattern as Figure 3 (youth report), except that it was based on data from the caregivers` report. Vertical dashed lines indicate the mean Δ brain embedding per group. Note that for the caregiver report there were not enough males in the 'postpubertal' category to allow for density plotting.

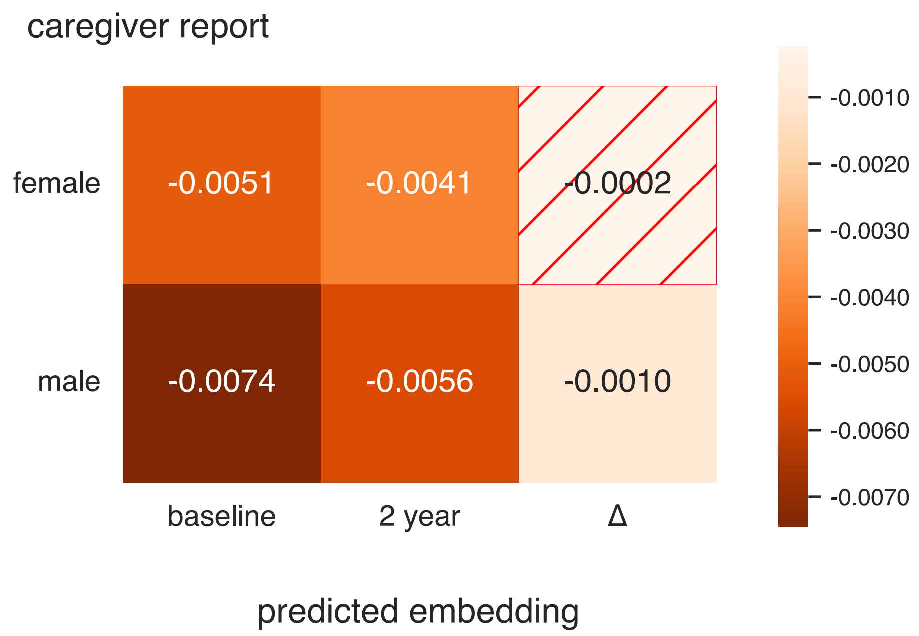

Supplementary Fig. 4: Associations between brain embeddings and dimensional psychopathology, both in cross-sectional and longitudinal data of the ABCD cohort. First two columns refer to associations between predicted embeddings and the respective psychopathology (CBCL Total) per timepoint. Δ refers to the association between the Δembedding and the ΔCBCL score. Annotations refer to effect sizes from linear models and hashed cells indicate non-significant results. Linear models were performed in similar vein to the puberty analyses in the main manuscript: cross-sectional models were calculated with age and site as covariate and the longitudinal model was complemented with site and Δage between both timepoints. Exact p-values and effect sizes can be found in Supplementary Table 1.

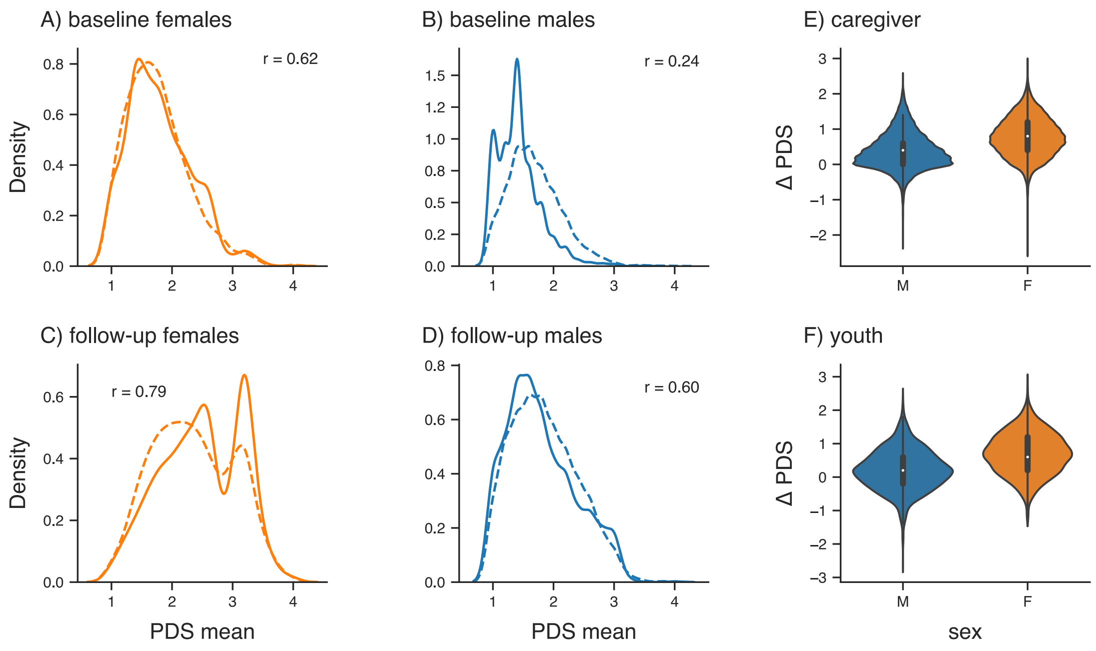

Supplementary Fig. 5: Distribution of puberty ratings in the ABCD sample stratified for sex, timepoints, and respondent (Panel A-D). Solid lines indicate caregiver and dashed line indicate youth reports. Higher correlation values between puberty ratings at follow-up indicate better alignment between reports from different sources. Panel E and F refer to changes in puberty scores (i.e., ΔPDS) for males and females. All plots only show data from participants for which data from both timepoints was available. r= Pearson correlation coefficient.

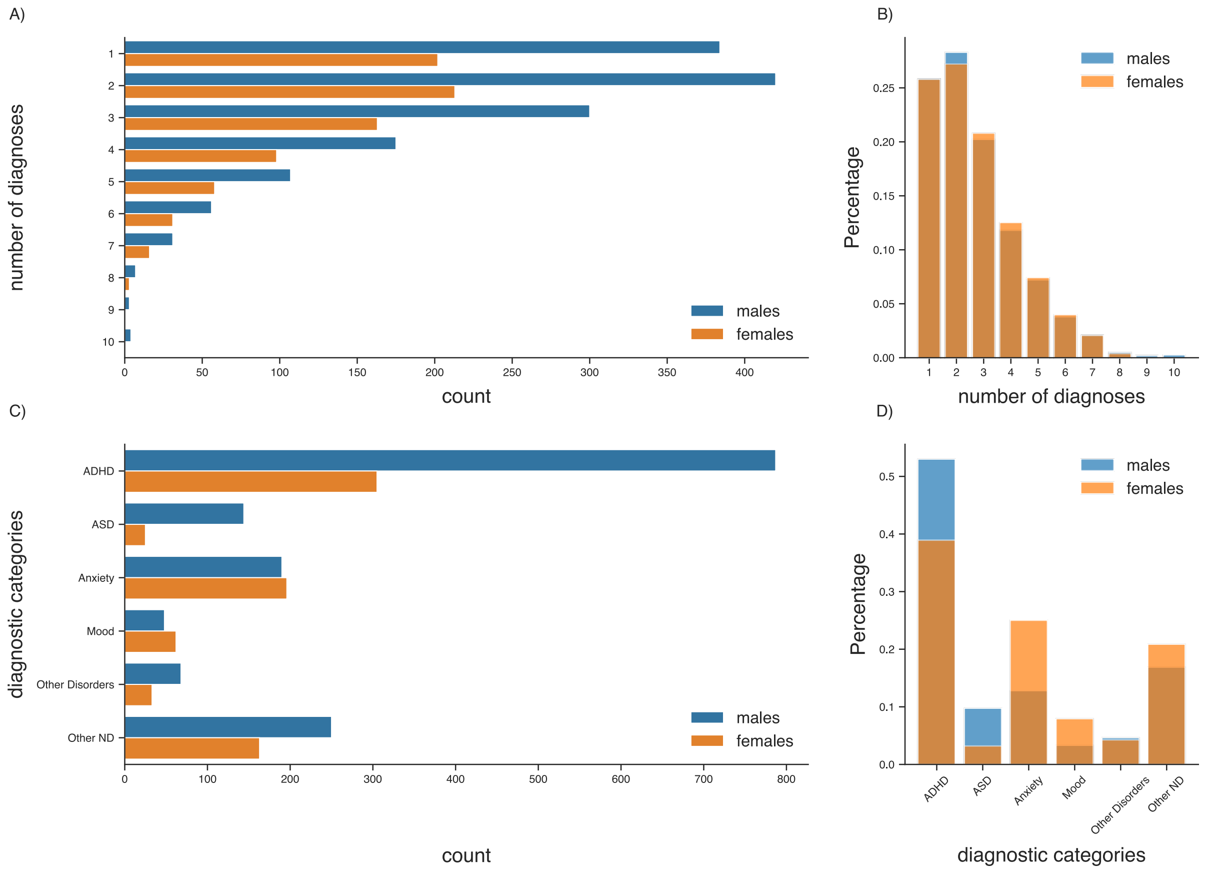

Supplementary Fig. 6: Distribution of psychopathology measures in the HBN sample as total counts and percentages stratified for sex. Diagnoses were grouped into six distinct categories following [3]. ADHD= Attention Deficit Hyperactivity Disorder, ASD= Autism Spectrum Disorder, ND= Neurodevelopmental Disorder

### Supplementary Tables

Supplementary Table 1: Association results for ABCD models.

| | sex | IV | b | p | $\eta^2$ |
|---|---|---|---|---|---|
| **Puberty** | | | | | |
| *cross-sectional* | | | | | |
| *embedding$_{baseline}$ ~ PDS + age + site* | | | | | |
| caregiver | female | PDS | -0.342 | $6.88 \times 10^{-16}$ | 0.021 |
| | | age | 0.001 | 0.623 | $7.00 \times 10^{-5}$ |
| | male | PDS | -0.358 | $5.53 \times 10^{-10}$ | 0.011 |
| | | age | -0.007 | 0.010 | 0.002 |

| | | | | | |
|---|---|---|---|---|---|
| youth | female | PDS | -0.173 | 0.005 | 0.006 |
| | | age | -0.003 | 0.435 | 0.0004 |
| | male | PDS | -0.057 | 0.338 | 0.0005 |
| | | age | -0.012 | 0.0005 | 0.006 |
| *embedding$_{2years}$ ~ PDS + age + site* | | | | | |
| caregiver | female | PDS | -0.270 | $1.94 \times 10^{-15}$ | 0.025 |
| | | age | -0.003 | 0.234 | 0.0004 |
| | male | PDS | -0.169 | $1.39 \times 10^{-5}$ | 0.008 |
| | | age | -0.006 | 0.029 | 0.001 |
| youth | female | PDS | -0.202 | $2.66 \times 10^{-9}$ | 0.015 |
| | | age | -0.004 | 0.161 | 0.0006 |
| | male | PDS | -0.141 | 0.0003 | 0.006 |
| | | age | -0.008 | 0.004 | 0.002 |
| *longitudinal* | | | | | |
| *Δembedding ~ ΔPDS + Δage + site* | | | | | |
| caregiver | female | ΔPDS | -0.058 | $2.37 \times 10^{-10}$ | 0.017 |
| | | Δage | -0.015 | $6.36 \times 10^{-11}$ | 0.014 |
| | male | ΔPDS | 0.012 | 0.191 | 0.0002 |
| | | Δage | -0.009 | $1.31 \times 10^{-5}$ | 0.005 |
| youth | female | ΔPDS | -0.083 | $3.79 \times 10^{-11}$ | 0.038 |
| | | Δage | -0.012 | 0.0004 | 0.009 |
| | males | ΔPDS | 0.009 | 0.358 | 0.0003 |

| | | | | | |
|---|---|---|---|---|---|
| | | Δage | -0.006 | 0.021 | 0.002 |
| *Δembedding ~ ΔPDS + $PDS_{baseline}$ + Δage + site* | | | | | |
| caregiver | females | ΔPDS | -0.083 | $7.88 \times 10^{-20}$ | 0.018 |
| | | $PDS_{baseline}$ | -0.138 | $1.10 \times 10^{-46}$ | 0.065 |
| | | Δage | -0.014 | $6.77 \times 10^{-11}$ | 0.014 |
| | males | ΔPDS | 0.010 | 0.289 | 0.0002 |
| | | $PDS_{baseline}$ | -0.015 | 0.238 | 0.0004 |
| | | Δage | -0.009 | $1.26 \times 10^{-5}$ | 0.005 |
| youth | females | ΔPDS | -0.113 | $2.38 \times 10^{-18}$ | 0.039 |
| | | $PDS_{baseline}$ | -0.120 | $1.15 \times 10^{-14}$ | 0.045 |
| | | Δage | -0.010 | 0.001 | 0.008 |
| | males | ΔPDS | 0.010 | 0.368 | 0.0003 |
| | | $PDS_{baseline}$ | 0.003 | 0.837 | $2.00 \times 10^{-5}$ |
| | | Δage | -0.006 | 0.021 | 0.002 |
| **Puberty** covarying for race/ethnicity, BMI, SES | | | | | |
| *Cross-sectional* | | | | | |
| *$embedding_{baseline}$ ~ PDS + age + site + BMI + SES + race/ethnicity* | | | | | |
| caregiver | females | PDS | -0.10 | 0.02 | 0.002 |
| | | SES | 0.19 | $4.81 \times 10^{-12}$ | 0.03 |
| | | BMI | 0.009 | 0.10 | 0.0 |
| | | age | -0.004 | 0.20 | 0.001 |
| | males | PDS | -0.01 | 0.84 | $1 \times 10^{-5}$ |
| | | SES | 0.23 | $1.65 \times 10^{-16}$ | 0.02 |
| | | BMI | 0.007 | 0.16 | $5 \times 10^{-5}$ |
| | | age | -0.009 | 0.001 | 0.003 |
| youth | females | PDS | -0.03 | 0.58 | 0.0002 |
| | | SES | 0.19 | $5.72 \times 10^{-6}$ | 0.015 |

| | | | | | |
|---|---|---|---|---|---|
| | | BMI | 0.01 | 0.15 | 0.0004 |
| | | age | -0.007 | 0.09 | 0.002 |
| | males | PDS | 0.03 | 0.62 | 0.0001 |
| | | SES | 0.17 | $1.47 \times 10^{-6}$ | 0.01 |
| | | BMI | 0.009 | 0.22 | 0.0002 |
| | | age | -0.01 | $9.45 \times 10^{-5}$ | 0.007 |
| *embedding$_{2years}$ ~ PDS + age + site + BMI + SES + race/ethnicity* | | | | | |
| caregiver | females | PDS | -0.09 | 0.02 | 0.002 |
| | | SES | 0.20 | $2.28 \times 10^{-11}$ | 0.02 |
| | | BMI | -0.002 | 0.73 | 0.002 |
| | | age | -0.008 | 0.01 | 0.006 |
| | males | PDS | -0.006 | 0.89 | $1 \times 10^{-5}$ |
| | | SES | 0.21 | $5.42 \times 10^{-12}$ | 0.01 |
| | | BMI | 0.0009 | 0.84 | 0.0002 |
| | | age | -0.01 | 0.0005 | 0.004 |
| youth | females | PDS | -0.07 | 0.059 | 0.001 |
| | | SES | 0.18 | $1.08 \times 10^{-8}$ | 0.01 |
| | | BMI | -0.006 | 0.20 | 0.003 |
| | | age | -0.007 | 0.032 | 0.004 |
| | males | PDS | 0.001 | 0.98 | 0.0 |
| | | SES | 0.20 | $2.28 \times 10^{-11}$ | 0.01 |
| | | BMI | 0.001 | 0.77 | 0.0001 |
| | | age | -0.01 | $2.35 \times 10^{-5}$ | 0.005 |
| *longitudinal* | | | | | |
| *Δembedding ~ ΔPDS + Δage + site ΔBMI + SES + race/ethnicity* | | | | | |
| caregiver | females | ΔPDS | -0.06 | $2.51 \times 10^{-9}$ | 0.014 |
| | | SES | 0.01 | 0.21 | 0.0004 |
| | | ΔBMI | 0.002 | 0.51 | 0.0 |
| | | Δage | -0.02 | $1.31 \times 10^{-7}$ | 0.012 |
| | males | ΔPDS | 0.01 | 0.38 | 0.0002 |

| | | | | | |
|---|---|---|---|---|---|
| | | SES | -0.006 | 0.43 | 0.0002 |
| | | ΔBMI | -0.003 | 0.19 | 0.0005 |
| | | Δage | -0.01 | 0.001 | 0.004 |
| youth | females | ΔPDS | -0.08 | $4.02 \times 10^{-8}$ | 0.03 |
| | | SES | 0.005 | 0.72 | 0.0 |
| | | ΔBMI | -0.005 | 0.15 | 0.003 |
| | | Δage | -0.01 | 0.02 | 0.008 |
| | males | ΔPDS | 0.004 | 0.71 | $8 \times 10^{-5}$ |
| | | SES | 0.006 | 0.52 | 0.0002 |
| | | ΔBMI | 0.0003 | 0.90 | 0.0 |
| | | Δage | -0.01 | 0.009 | 0.004 |
| *Δembedding ~ ΔPDS + Δage + site ΔBMI + $BMI_{baseline}$ + SES + race/ethnicity* | | | | | |
| caregiver | females | ΔPDS | -0.06 | $9.60 \times 10^{-9}$ | 0.013 |
| | | SES | 0.005 | 0.56 | $4 \times 10^{-5}$ |
| | | ΔBMI | 0.003 | 0.26 | 0.0001 |
| | | BMI | -0.01 | $1.70 \times 10^{-7}$ | 0.012 |
| | | Δage | -0.02 | $4.75 \times 10^{-8}$ | 0.012 |
| | males | ΔPDS | 0.01 | 0.21 | 0.0005 |
| | | SES | -0.009 | 0.23 | 0.0004 |
| | | ΔBMI | -0.002 | 0.23 | 0.0004 |
| | | BMI | -0.005 | 0.0003 | 0.0039 |
| | | Δage | -0.01 | 0.001 | 0.0036 |
| youth | females | ΔPDS | -0.08 | $1.18 \times 10^{-7}$ | 0.026 |
| | | SES | -0.001 | 0.93 | 0.0002 |
| | | ΔBMI | -0.004 | 0.22 | 0.002 |
| | | BMI | -0.007 | 0.002 | 0.012 |
| | | Δage | -0.01 | 0.02 | 0.008 |
| | males | ΔPDS | 0.009 | 0.37 | 0.0004 |
| | | SES | 0.002 | 0.85 | $2 \times 10^{-5}$ |
| | | ΔBMI | 0.001 | 0.64 | 0.0001 |

| | | | | | |
|---|---|---|---|---|---|
| | | BMI | -0.007 | $1.67 \times 10^{-5}$ | 0.01 |
| | | Δage | -0.01 | 0.008 | 0.004 |
| *Δembedding ~ ΔPDS + $PDS_{baseline}$ + Δage + site ΔBMI + $BMI_{baseline}$ + SES + race/ethnicity* | | | | | |
| caregiver | females | ΔPDS | -0.08 | $1.49 \times 10^{-15}$ | 0.024 |
| | | PDS | -0.13 | $2.06 \times 10^{-25}$ | 0.030 |
| | | SES | -0.004 | 0.63 | $4 \times 10^{-5}$ |
| | | ΔBMI | 0.006 | 0.008 | 0.0001 |
| | | BMI | -0.002 | 0.29 | 0.012 |
| | | Δage | -0.02 | $1.61 \times 10^{-7}$ | 0.013 |
| | males | ΔPDS | 0.01 | 0.24 | 0.0005 |
| | | PDS | -0.004 | 0.78 | 0.0001 |
| | | SES | -0.009 | 0.23 | 0.0004 |
| | | ΔBMI | -0.002 | 0.23 | 0.0004 |
| | | BMI | -0.004 | 0.0004 | 0.004 |
| | | Δage | -0.01 | 0.0007 | 0.004 |
| youth | females | ΔPDS | -0.10 | $3.78 \times 10^{-11}$ | 0.04 |
| | | PDS | -0.09 | $1.66 \times 10\text{-}^{6}$ | 0.007 |
| | | SES | -0.006 | 0.65 | 0.0002 |
| | | ΔBMI | -0.003 | 0.49 | 0.0025 |
| | | BMI | -0.003 | 0.15 | 0.013 |
| | | Δage | -0-01 | 0.04 | 0.0086 |
| | males | ΔPDS | 0.01 | 0.24 | 0.0008 |
| | | PDS | 0.01 | 0.41 | $3 \times 10^{-5}$ |
| | | SES | 0.002 | 0.83 | $2 \times 10^{-5}$ |
| | | ΔBMI | 0.001 | 0.63 | 0.0001 |
| | | BMI | -0.008 | $1.20 \times 10^{-5}$ | 0.01 |
| | | Δage | -0.01 | 0.008 | 0.0038 |
| **psychopathology** | | | | | |
| *$embedding_{baseline}$ ~ CBCL_Total + age + site* | | | | | |
| caregiver | females | CBCL | -0.005 | $2.86 \times 10^{-5}$ | 0.005 |

| | | | | | |
|---|---|---|---|---|---|
| | | age | -0.004 | 0.10 | 0.0008 |
| | males | CBCL | -0.007 | $8.42 \times 10^{-8}$ | 0.01 |
| | | age | -0.009 | 0.001 | 0.003 |
| *Embedding2years ~ CBCL_Total + age + site* | | | | | |
| caregiver | females | CBCL | -0.004 | 0.002 | 0.004 |
| | | age | -0.001 | 0.002 | 0.003 |
| | males | CBCL | -0.006 | $2.34 \times 10^{-6}$ | 0.006 |
| | | age | -0.009 | 0.002 | 0.003 |
| *Δembedding ~ ΔCBCL + Δage + site* | | | | | |
| caregiver | females | ΔCBCL | -0.002 | 0.50 | 0.0002 |
| | | Δage | -0.17 | $1.73 \times 10^{-9}$ | 0.01 |
| | males | ΔCBCL | -0.001 | 0.005 | 0.002 |
| | | Δage | -0.01 | 0.0001 | 0.004 |

Note: All models included scan site as covariate of no interest, thus site is not included in the table above for clarity. Additionally race/ethnicity is also not included for clarity. Bonferroni-corrected $\alpha_{puberty}$: 0.05 / 12 = 0.004, $\alpha_{psychopathology}$: 0.05 / 6 = 0.008. IV = independent variable, $\eta^2$ = partial eta squared, CBCL= Child Behavior Check List.

Δ refers to differences in the respective variable between baseline and 2-years follow up.

Supplementary Table 2: Association results for HBN models.

| | **sex** | **IV** | **b** | **p** | $\eta^2$ |
|---|---|---|---|---|---|
| **psychopathology** sum of diagnoses | | | | | |
| *embedding ~ sum of diagnoses + age + site* | | | | | |
| | female | $diagnoses_{sum}$ | .07 | .007 | .007 |
| | | age | -.02 | .08 | .004 |
| | male | $diagnoses_{sum}$ | .04 | .05 | .001 |
| | | age | -.051 | $1.04 \times 10^{-8}$ | .022 |

| | | | | | |
|---|---|---|---|---|---|
| *embedding ~ sum of diagnoses + PDS + age + site* | | | | | |
| | female | $diagnoses_{sum}$ | .074 | .01 | .010 |
| | | PDS | .166 | .07 | $3.00 \times 10^{-5}$ |
| | | age | -.057 | .012 | .011 |
| | male | $diagnoses_{sum}$ | .037 | .101 | .002 |
| | | PDS | .051 | .499 | .010 |
| | | age | -.061 | .0008 | .010 |
| *embedding ~ sum of diagnoses + PDS + diag:PDS + age + site* | | | | | |
| | females | $diagnoses_{sum}$ | .10 | .19 | .010 |
| | | PDS | .19 | .13 | $3.00 \times 10^{-5}$ |
| | | diag:PDS | -.01 | .75 | .0005 |
| | | age | -.056 | .014 | .010 |
| | male | $diagnoses_{sum}$ | .012 | .819 | .002 |
| | | PDS | .012 | .912 | .010 |
| | | diag:PDS | .013 | .598 | .0003 |
| | | age | -.061 | .0008 | .010 |
| **psychopathology** CBCL Total Score | | | | | |
| *embedding ~ CBCL_Total + age + site* | | | | | |
| | female | CBCL | 0.004 | 0.008 | 0.01 |
| | | age | -0.007 | 0.59 | 0.0005 |
| | male | CBCL | 0.002 | 0.06 | 0.004 |
| | | age | -0.05 | $5.98 \times 10^{-7}$ | 0.02 |

| | | | | | |
|---|---|---|---|---|---|
| *embedding ~ CBCL_Total + PDS + age + site* | | | | | |
| | female | CBCL | 0.005 | 0.002 | 0.02 |
| | | PDS | 0.22 | 0.04 | 0.001 |
| | | age | -0.06 | 0.04 | 0.01 |
| | male | CBCL | 0.001 | 0.39 | 0.001 |
| | | PDS | -0.01 | 0.88 | 0.01 |
| | | age | -0.05 | 0.014 | 0.006 |
| *embedding ~ CBCL_Total + PDS + CBCL:PDS + age + site* | | | | | |
| | female | CBCL | 0.008 | 0.08 | 0.02 |
| | | PDS | 0.27 | 0.05 | 0.001 |
| | | CBCL:PDS | -0.001 | 0.57 | 0.0009 |
| | | age | -0.06 | 0.04 | 0.001 |
| | male | CBCL | 0.004 | 0.32 | 0.001 |
| | | PDS | 0.04 | 0.71 | 0.01 |
| | | CBCL:PDS | -0.001 | 0.49 | 0.0004 |
| | | age | -0.05 | 0.01 | 0.007 |
| **puberty** age restricted sample* matching ABCD age range at baseline and follow-up<br>*baseline: n=141 females, n=300 males, age: 8.92 – 11.08 years<br>follow-up: n=134 females, n=259 males, age: 10.59 – 13.38 years | | | | | |
| *embedding ~ PDS + $age_{baseline\text{-}matched}$ + site* | | | | | |
| | female | PDS | 0.33 | 0.07 | 0.02 |
| | | age | -0.05 | 0.76 | 0.0007 |

| | | | | | |
|---|---|---|---|---|---|
| | male | PDS | 0.07 | 0.68 | 0.003 |
| | | age | 0.20 | 0.07 | 0.01 |
| *embedding ~ PDS + age<sub>follow-up-matched</sub> + site* | | | | | |
| | female | PDS | -0.01 | 0.95 | 0.0018 |
| | | age | 0.11 | 0.26 | 0.01 |
| | male | PDS | -0.12 | 0.37 | 0.015 |
| | | age | -0.17 | 0.07 | 0.013 |

Note: All models included scan site as covariate of no interest, thus site is not included in the table above for clarity. Bonferroni-corrected $\alpha$: 0.05 / 2 = 0.025, IV = independent variable, $\eta^2$ = partial eta squared. CBCL = Child Behavior Checklist. Diag/CBCL:PDS refers to interaction term between $\text{diagnosis}_{\text{sum}}$ / CBCL and PDS.

Supplementary Table 3: Pubertal category conversion scheme

| sex | conversion scheme | pubertal category |
|---|---|---|
| female | premenarcheal + pubertal score = 2 | prepubertal |
| | premenarcheal + pubertal score = 3 | early pubertal |
| | premenarcheal + pubertal score >= 3 | mid pubertal |
| | postmenarcheal + pubertal score <= 7 | late pubertal |
| | postmenarcheal + pubertal score = 8 | postpubertal |
| male | pubertal score = 3 | prepubertal |
| | pubertal score >= 4 and <=5 | early pubertal |
| | pubertal score >= 6 and <=8 | mid pubertal |
| | pubertal score >= 9 and <=11 | late pubertal |

| | pubertal score = 12 | postpubertal |
|---|---|---|

Note: Pubertal score refers to the Pubertal Category Score, which depicts the sum of three/two PDS items for male and female, respectively. Male: pubic + facial hair growth + voice deepening. Female: pubic hair growth + breast development. Conversion follows the ABCD variables *'pds_p_ss_female_category'* and *'pds_p_ss_male_category'.*

Supplementary Table 4: Frequencies of pubertal categories in the ABCD sample.

| Pubertal Category | Caregiver Report | | Youth Report | |
|---|---|---|---|---|
| *baseline visit* | | | | |
| | male (N= 4045) | female (N= 3487) | male (N= 3761) | female (N= 2562) |
| prepubertal | n= 2907 (71,9%) | n= 1129 (32,4%) | n= 1134 (30,2%) | n= 679 (26,5%) |
| early pubertal | n= 932 (23.0%) | n= 818 (23,5%) | n= 1783 (47,4%) | n= 697 (27,2%) |
| mid pubertal | n= 184 (4.5%) | n= 1441(41,3%) | n= 779 (20,7%) | n= 1097 (42,8%) |
| late pubertal | n= 20 (<1%) | n= 86 (2%) | n= 57 (1,5%) | n= 82 (3%) |
| postpubertal | n= 2 (<1%) | n= 4 (<1%) | n= 8 (<1%) | n= 7 (<1%) |
| *2 years follow up visit* | | | | |
| | male (N=3970) | female (N= 3378) | male (N= 4095) | female (N= 3299) |

| | | | | |
|---|---|---|---|---|
| prepubertal | n= 1448 (36,5%) | n= 147 (4,4%) | n= 878 (21,4%) | n= 162 (4,9%) |
| early pubertal | n= 1490 (37,5%) | n= 323 (9,5%) | n= 1715 (41,9%) | n= 360 (10,9%) |
| mid pubertal | n= 834 (21%) | n= 1601 (47,4%) | n= 1342 (32,8%) | n= 1563 (47,4%) |
| late pubertal | n= 194 (4,9%) | n= 1249 (37%) | n= 155 (3,8%) | n= 1166 (35,3%) |
| postpubertal | n= 4 (<1%) | n= 58 (1,7%) | n= 5 (<1%) | n= 48 (1,5%) |

Supplementary Table 5: Frequencies of psychopathology measures in the HBN sample.

| | male (N=1487) | female (N=784) |
|---|---|---|
| *Number of diagnoses* | | |
| 1 | n= 384 (25,8%) | n= 202 (25,8%) |
| 2 | n= 420 (28,2%) | n= 231 (27,2%) |
| 3 | n= 300 (20,2%) | n= 163 (20,8%) |
| 4 | n= 175 (11,8%) | n= 98 (12,5%) |
| 5 | n= 107 (7,2%) | n= 58 (7,4%) |
| 6 | n= 56 (3,8%) | n= 31 (4,0%) |
| 7 | n= 31 (2,1%) | n= 16 (2,0%) |
| 8 | n= 7 (< 1%) | n= 3 (< 1%) |
| 9 | n= 3 (< 1%) | - |
| 10 | n= 4 (< 1%) | - |
| mean (SD) | 2.71 (1.62) | 2.71 (1.55) |
| *Primary Diagnosis* | | |
| ADHD | n= 787 (52,9%) | n= 305 (38,9%) |

| | | |
|---|---|---|
| ASD | n= 144 (9,7%) | n= 25 (3,2%) |
| Anxiety | n= 190 (12,8%) | n= 196 (25,0%) |
| Mood | n= 48 (3,2%) | n= 62 (7,9%) |
| Other Disorder | n= 68 (4,6%) | n= 33 (4,2%) |
| Other ND | n= 250 (16,8%) | n= 163 (20,8%) |
| *CBCL Total* | n= 1269 | n= 635 |
| mean (SD) | 43,93 (25,99) | 42,60 (25,39) |

Note: ADHD= Attention Deficit Hyperactivity Disorder, ASD= Autism Spectrum Disorder, ND= Neurodevelopmental Disorder; SD = standard deviation.

**Supplementary References**

1. Mowinckel, A. M. & Vidal-Piñeiro, D. Visualization of Brain Statistics With R Packages *ggseg* and *ggseg3d*. *Adv. Methods Pract. Psychol. Sci.* 3, 466–483 (2020).
2. Larivière, S. *et al.* The ENIGMA Toolbox: multiscale neural contextualization of multisite neuroimaging datasets. *Nat. Methods* 18, 698–700 (2021).
3. Voldsbekk, I. *et al. Delineating disorder-general and disorder-specific dimensions of psychopathology from functional brain networks in a developmental clinical sample.* http://medrxiv.org/lookup/doi/10.1101/2023.03.31.23288009 (2023) doi:10.1101/2023.03.31.23288009.
